# Evaluation of the diagnostic value of YiDiXie™-SS, YiDiXie™-HS and YiDiXie™-D in renal cancer

**DOI:** 10.1101/2024.07.28.24310613

**Authors:** Yutong Wu, Wenkang Chen, Yingqi Li, Chen Sun, Zhenjian Ge, Shengjie Lin, Pengwu Zhang, Wuping Wang, Siwei Chen, Huimei Zhou, Xutai Li, Wei Li, Xionghui Wu, Liangkuan Bi, Yongqing Lai

## Abstract

**Background:** Renal cancer is a serious threat to human health and causes heavy economic burden. Enhanced CT is widely used in the diagnosis of renal tumors. However, false-positive results on enhanced CT can lead to misdiagnosis and incorrect surgery or treatment, while false-negative results on enhanced CT can lead to missed diagnosis and delayed treatment. There is an urgent need to find convenient, cost-effective and non-invasive diagnostic methods to reduce the false-positive and false-negative rates of enhanced CT in renal tumors. The aim of this study was to evaluate the diagnostic value of YiDiXie™-SS, YiDiXie™-HS and YiDiXie™-D in renal cancer.

**Patients and methods:** 309 subjects (malignant group, n=244; benign group, n=65) were finally included in this study. Remaining serum samples from the subjects were collected and tested by applying the YiDiXie™all-cancer detection kit to evaluate the sensitivity and specificity of YiDiXie™-SS, YiDiXie™-HS and YiDiXie™-D, respectively.

**Results:** The sensitivity of YiDiXie™SS was 98.4% (95.9% - 99.4%) and its specificity was 70.8% (58.8% - 80.4%). This means that YiDiXie™-SS has very high sensitivity and specificity in renal tumors.YiDiXie™-HS has a sensitivity of 85.2% (80.3% - 89.1%) and its specificity is 84.6% (73.9% - 91.4%). This means that YiDiXie™-HS has both the sensitivity and specificity in kidney tumors.YiDiXie™-D has a sensitivity of 72.5% (66.6% - 77.8%) and its specificity is 92.3% (83.2% - 96.7%). This means that YiDiXie™-D has high sensitivity and very high specificity in kidney tumors.YiDiXie™-SS had a sensitivity of 98.6% (95% CI: 95.8% - 99.6%) and a specificity of 71.4% (95% CI: 45.4% - 88.3%) in renal enhanced CT-positive patients. This means that the application of YiDiXie™-SS reduces the false-positive rate of renal enhanced CT by 71.4% (95% CI: 45.4% - 88.3%) with essentially no increase in malignant tumor leakage. The sensitivity of YiDiXie ™-HS in renal enhanced CT-negative patients was 86.5% (95% CI: 72.0% - 94.1%) and its specificity was 84.3% (95% CI: 72.0% - 91.8%). This means that the application of YiDiXie™-HS reduces the false-negative rate of enhanced CT by 86.5% (95% CI: 72.0% - 94.1%).The sensitivity of YiDiXie™-D in renal enhanced CT-positive patients was 31.9% (95% CI: 25.9% - 38.5%) and its specificity was 92.9% (95% CI: 68.5% - 99.6%). This means that YiDiXie™-D reduces the false positive rate of enhanced CT by 92.9% (95% CI: 68.5% - 99.6%). YiDiXie™-D had a sensitivity of 64.9% (95% CI: 48.8% - 78.2%) and a specificity of 92.2% (95% CI: 81.5% - 96.9%) in patients with negative enhanced CT. This means that YiDiXie™-D reduces the false-negative rate of enhanced CT by 64.9% (95% CI: 48.8% - 78.2%) while maintaining high specificity.

**Conclusion:** YiDiXie™-SS has very high sensitivity and high specificity in renal tumors. YiDiXie™-HS has high sensitivity and high specificity in renal tumors. YiDiXie™-D has high sensitivity and very high specificity in renal tumors. YiDiXie™-SS significantly reduces renal enhancement CT false-positive rates with essentially no increase in delayed treatment of malignant tumors. YiDiXie™-HS significantly reduces the false-negative rate of renal enhancement CT. YiDiXie™-D can significantly reduce the false-positive rate of renal enhancement CT or significantly reduce the false-negative rate while maintaining high specificity. the YiDiXie™test has important diagnostic value in renal cancer and is expected to solve the two problems of “too high false-positive rate” and “too high false-negative rate” of renal enhanced CT.

**Clinical trial number:** ChiCTR2200066840.

## INTRODUCTION

Renal cancer is one of the common malignant tumors of the urinary system^1^. The latest data show that there are 434,419 new kidney cancer cases and 155,702 new deaths worldwide in 2022^2^; compared with 2020, the incidence rate of RCC increased by 0.7% in 2022^3^, which is still showing a trend of increasing year by year. Since renal cancer is insensitive and less effective to treatments such as radiotherapy and chemotherapy, and is prone to metastasis and recurrence, surgical resection is still considered the most effective treatment for limited renal cancer without metastasis^4^. Renal cancer has an insidious onset with no specific clinical manifestations in the early stage, and about 33% of patients have already developed distant metastasis at the time of the first visit^5^, and 20%-30% of patients develop recurrence at postoperative follow-up^6^. Studies have shown that the 5-year survival rate for stage I kidney cancer can be as high as 84%-90%, compared to only 6% for stage IV kidney cancer^7^. Through early screening, not only the prognosis of patients can be improved, but also the economic burden of patients can be reduced^8^. Thus, kidney cancer is a serious threat to human health.

Enhanced CT is widely used in the diagnosis of renal tumors. On the one hand, enhanced CT can produce a large number of false-positive results. The false-positive rate of enhanced CT in smaller renal tumors can reach 15-30%^9,10^. When enhancement CT is positive, patients usually undergo tumor resection or radical resection^4^. A false-positive result on enhanced CT means that a benign disease is misdiagnosed as a malignant tumor, and the patient will have to bear the undesirable consequences of unnecessary mental anguish, costly surgeries and investigations, physical injuries, and even organ removal and loss of function. Therefore, there is an urgent need to find a convenient, cost-effective and noninvasive diagnostic method to reduce the false-positive rate of enhanced CT for renal tumors.

On the other hand, enhanced CT can produce a large number of false negative results. The sensitivity of enhanced CT for diagnosing renal cancer is less than 80%^11^. The imaging characteristics of enhanced CT make it less accurate in evaluating renal tumors <5 cm, with a false-negative rate of 28%^12^. When enhancement CT is negative, patients are usually taken to observation, with regular follow-up^4^. Its false-negative result means that the malignancy has been missed and will likely lead to a delay in treatment, progression of the malignancy, and possibly even development of an advanced stage. Patients will thus have to bear the adverse consequences of poor prognosis, high treatment costs, poor quality of life, and short survival. Therefore, there is an urgent need to find a convenient, economical and noninvasive diagnostic method to reduce the false-negative rate of enhanced CT for renal tumors.

In addition, there are some special patients who need to be extra cautious in choosing whether or not to operate, such as: smaller tumors, tumors with difficulty in preserving the kidney, tumors requiring radical surgery, tumors in isolated kidneys, contralateral renal insufficiency, total renal insufficiency, and patients with poor general conditions. The risk of wrong surgery in these special patients is much higher than the risk of missed diagnosis. And false-positive results mean that benign diseases are misdiagnosed as malignant tumors, which will lead to wrong diagnosis and wrong surgery. Therefore, there is an urgent need to find a convenient, cost-effective and noninvasive diagnostic method with high specificity to substantially reduce the false-positive rate of renal enhancement CT in these special patients or to significantly reduce its false-negative rate while maintaining high specificity.

Based on the detection of novel tumor markers of miRNA in serum, Shenzhen KeRuiDa Health Technology Co., Ltd. has developed an in vitro diagnostic test, YiDiXie ™all-cancer test (hereinafter referred to as YiDiXie™test), which can detect multiple types of cancers with only 200 microliters of whole blood or 100 microliters of serum each time^13^. The YiDiXie ™test consists of three different tests, YiDiXie™-HS, YiDiXie™-SS and YiDiXie™-D^13^.

The purpose of this study was to evaluate the diagnostic value of three tests of the YiDiXie™test, YiDiXie ™-SS, YiDiXie ™-HS, and YiDiXie ™-D, in renal cancer.

## PATIENTS AND METHODS

### Study design

This work is part of the sub-study “Evaluating the diagnostic value of the YiDiXie ™test in multiple tumors” of the SZ-PILOT study (ChiCTR2200066840).

The SZ-PILOT study (ChiCTR2200066840) is a single-center, prospective, observational study. Participants who signed a general informed consent for donation of remaining samples at the time of admission or physical examination were included, and whose remaining serum samples of 0.5 ml were collected for use in this study.

The study was blinded. Both the laboratory personnel who performed the YiDiXie ™test and the KeRuiDa laboratory technicians who adjudicated the results of the YiDiXie ™test were unaware of the subjects’ clinical information. The clinical experts who evaluated the subjects’ clinical information were also unaware of the results of the YiDiXie™test.

The study was approved by the Ethics Committee of Peking University Shenzhen Hospital and was conducted in accordance with the International Conference on Harmonization for “Good clinical practice guidelines” and the Declaration of Helsinki.

### Participants

Subjects with positive renal ultrasound were included in this study. The two groups of participants were enrolled separately and all participants who met the inclusion criteria were included consecutively.

The present study initially included hospitalized patients with “suspected (solid or hematological) malignancy” who signed a general informed consent for donation of the remaining samples. Participants with a postoperative pathological diagnosis of “malignant tumor” were included in the malignant group, and those with a postoperative pathological diagnosis of “benign tumor” were included in the benign group. Subjects with ambiguous pathologic results were excluded from the study. Some of the samples from the malignant group were used in our previous work^13^.

Participants failing the serum sample quality test prior to the YiDiXie™test were excluded from this study. For details of enrollment and exclusion, please refer to our previous work^13^.

### Sample collection, processing

The serum samples used in this study were obtained from serum left over after a normal clinic visit, without the need for additional blood draws. Approximately 0.5 ml of serum samples were collected from the remaining serum of subjects in the Medical Laboratory and stored at -80°C for use in the subsequent YiDiXie™test.

### The YiDiXie™test

The YiDiXie ™test is performed through the YiDiXie ™all-cancer detection kit, an in-vitro diagnostic kit developed and manufactured by Shenzhen KeRuiDa Health Technology Co.. The kit detects the expression levels of dozens of miRNA biomarkers in the serum to judge the presence or absence of cancer in the subject’s body. An appropriate threshold is predefined for each miRNA biomarker to ensure that each miRNA marker is highly specific, and these independent assays are integrated in a concurrent assay format to dramatically increase the sensitivity of broad-spectrum cancers and maintain high specificity.

The YiDiXie ™test consists of three distinct assays: YiDiXie™-HS, YiDiXie™-SS, and YiDiXie™-D. YiDiXie™-Highly Sensitive (YiDiXie™-HS) has been developed with a balance of sensitivity and specificity. YiDiXie™-Super Sensitive (YiDiXie™-SS) significantly increases the number of miRNA tests to achieve extremely high sensitivity for all clinical stages of all malignant tumor types. YiDiXie ™-Diagnosis (YiDiXie ™-D) drastically enhances the diagnostic thresholds of individual miRNA tests to achieve extremely high specificity for all malignant tumor types.

The YiDiXie ™test was conducted following the instructions for the YiDiXie ™all-cancer detection kit. Detailed procedures are described in our previous work^13^.

The original results were analyzed by the laboratory technicians of Shenzhen KeRuiDa Health Technology Co., Ltd and the results of the YiDiXie™test were determined to be “positive” or “negative”.

### Diagnosis of Enhanced CT

The diagnostic conclusion of the enhanced CT is judged to be “positive” or “negative”. If the diagnostic conclusion is positive, relatively positive, or favors a malignant tumor, the test result is considered “positive”. If the diagnosis is positive, more positive, or favors a benign tumor, or if the diagnosis is ambiguous, the result is considered “negative”.

### Extraction of clinical data

The clinical, pathological, laboratory, and imaging data included in this study were drawn from the subjects’ hospitalized medical records or physical examination reports. The clinical staging was completed by trained clinicians assessed according to the AJCC staging manual (7th or 8th edition)^17,18^.

### Statistical analyses

Descriptive statistics were reported for demographic and baseline characteristics. For categorical variables, the number and percentage of subjects in each category were calculated; for continuous variables, the total number of subjects (n), mean, standard deviation (SD) or standard error (SE), median, first quartile (Q1), third quartile (Q3), minimum, and maximum values were calculated. 95% confidence intervals (CI) for multiple indicators were calculated using the Wilson (score) method.

## RESULTS

### Participant disposition

309 participants were finally included in this study (malignant group, n=244; benign group, n=65). The demographic and clinical characteristics of the 309 participants in the study are presented in Table 1.

**Table 1.** Participants’ demographic and clinical manifestation.

|  | Cancer (n = 244) | Benign (n = 65) | Total (N = 309) |
| --- | --- | --- | --- |
| Age, years |  |  |  |
| Mean (SD) | 53.1 (12.75) | 44.2 (12.76) | 51.2 (13.26) |
| Median (Q1,Q3) | 54 (44, 64) | 42 (35, 54) | 51 (41, 62) |
| Min, max | 19, 83 | 19, 68 | 19, 83 |
| Age, group, n (%) |  |  |  |
| < 50 | 95 (38.9) | 41 (63.1) | 136 (44.0) |
| ≥ 50 | 149 (61.1) | 24 (36.9) | 173 (56.0) |
| < 65 | 189 (77.5) | 61 (93.8) | 250 (80.9) |
| ≥ 65 | 55 (22.5) | 4 (6.2) | 59 (19.1) |
| Sex, n (%) |  |  |  |
| Female | 77 (31.6) | 53 (81.5) | 130 (42.1) |
| Male | 167 (68.4) | 12 (18.5) | 179 (57.9) |
| Body mass index (kg/m2) |  |  |  |
| n | 150 | 61 | 211 |
| Mean (SD) | 24.3 (3.71) | 22.6 (3.48) | 23.8 (3.71) |
| Median (Q1,Q3) | 24.2 (21.5, 26.1) | 22.3 (20.1, 25.0) | 23.7 (20.7, 25.8) |
| Min, max | 16.4 39.7 | 17.8 31.2 | 16.4 39.7 |
| Body mass index category, n (%) |  |  |  |
| Underweight | 4 (1.6) | 6 (9.2) | 10 (3.2) |
| Benign | 68 (27.9) | 35 (53.8) | 103 (33.3) |
| Overweight | 55 (22.5) | 15 (23.1) | 70 (22.7) |
| Obese | 23 (9.4) | 5 (7.7) | 28 (9.1) |
| Missing | 94 (38.5) | 4 (6.2) | 98 (31.7) |
| AJCC clinical stage |  |  |  |
| Stage I | 199 (81.6) |  | 199 (81.6) |
| Stage II | 34 (13.9) |  | 34 (13.9) |
| Stage III | 11 (4.5) |  | 11 (4.5) |

Q1,Q3, first quartile, third quartile; SD, standard deviation.

The two groups of study subjects were comparable in terms of demographic and clinical characteristics (Table 1). The mean (standard deviation) age was 51.2(13.26)years and 42.1%(130/309) were female.

### Diagnostic performance of YiDiXie™-SS

As shown in Table 2, the sensitivity of YiDiXie™SS was 98.4% (95% CI: 95.9% - 99.4%) and its specificity was 70.8% (95% CI: 58.8% - 80.4%). This means that YiDiXie ™SS has very high sensitivity and high specificity in kidney tumors.

**Table 2.** Performance of YiDiXie™-SS.

|  | Malignant | Benign | Total |
| --- | --- | --- | --- |
|  | 244 | 65 | 309 |
| Positive | 240 | 19 | 259 |
| Negative | 4 | 46 | 50 |

SEN, Sensitivity. SPE, Specificity. Two-sided 95% Wilson confidence intervals were calculated.

### Diagnostic performance of YiDiXie™-HS

As shown in Table 3, the sensitivity of YiDiXie™-HS was 85.2% (95% CI: 80.3% - 89.1%) and its specificity was 84.6% (95% CI: 73.9% - 91.4%). This means that YiDiXie ™-HS has high sensitivity and high specificity in kidney tumors.

**Table 3.** Performance of YiDiXie™-HS.

|  | Malignant | Benign | Total |
| --- | --- | --- | --- |
|  | 244 | 65 | 309 |
| Positive | 208 | 10 | 218 |
| Negative | 36 | 55 | 91 |

SEN, Sensitivity. SPE, Specificity. Two-sided 95% Wilson confidence intervals were calculated.

### Diagnostic performance of YiDiXie™-D

As shown in Table 4, the sensitivity of YiDiXie™-D was 72.5% (95% CI: 66.6% - 77.8%) and its specificity was 92.3% (95% CI: 83.2% - 96.7%). This means that YiDiXie ™-D has high sensitivity and very high specificity in kidney tumors.

**Table 4.** Performance of YiDiXie™-D.

|  | Malignant | Benign | Total |
| --- | --- | --- | --- |
|  | 244 | 65 | 309 |
| Positive | 177 | 5 | 182 |
| Negative | 67 | 60 | 127 |

SEN = $177/244 = 72.5\%$ (66.6% - 77.8%) SPE = $60/65 = 92.3\%$ (83.2% - 96.7%)
SEN, Sensitivity. SPE, Specificity. Two-sided 95% Wilson confidence intervals were calculated.

### Diagnostic performance of enhanced CT

As shown in Table 5, the sensitivity of enhanced CT was 84.8%(95% CI: 79.8% - 88.8%) and its specificity was 78.5%(95% CI: 67.0% - 86.7%).

**Table 5.** Performance of different Items.

| Items | Total N | Positive | Sensitivity % (95% CI)<sup>a</sup> | Total N | Negative | Specificity % (95% CI)<sup>b</sup> |
| --- | --- | --- | --- | --- | --- | --- |
| CT | 244 | 207 | 84.8% (79.8% - 88.8%) | 65 | 51 | 78.5% (67.0% - 86.7%) |
| YiDiXie™-SS in CT-positive | 207 | 204 | 98.6% (95.8% - 99.6%) | 14 | 10 | 71.4% (45.4% - 88.3%) |
| YiDiXie™-HS in CT-negative | 37 | 32 | 86.5% (72.0% - 94.1%) | 51 | 43 | 84.3% (72.0% - 91.8%) |
| YiDiXie™-D in CT-positive | 207 | 153 | 73.9% (67.5% - 79.4%) | 14 | 13 | 92.9% (68.5% - 99.6%) |
| YiDiXie™-D in CT-negative | 37 | 24 | 64.9% (48.8% - 78.2%) | 51 | 47 | 92.2% (81.5% - 96.9%) |

CI, confidence interval. <sup>a,b</sup>Two-sided 95% Wilson confidence intervals were calculated.

### Diagnostic performance of YiDiXie™-SS in renal enhanced CT-positive patients

In order to solve the challenge of high false-positive rate of renal-enhanced CT, YiDiXie™-SS was applied to renal-enhanced CT-positive patients.

As shown in Table 5, the sensitivity of YiDiXie™-SS in patients with positive renal enhancement CT was 98.6% (95% CI: 95.8% - 99.6%) and its specificity was 71.4% (95% CI: 45.4% - 88.3%). This means that the application of YiDiXie ™SS reduces the false-positive rate of renal enhancement CT by 71.4% (95% CI: 45.4% - 88.3%) with essentially no increase in malignant tumor underdiagnosis.

### Diagnostic Performance of YiDiXie™-HS in renal enhanced CT-negative patients

In order to solve the challenge of high leakage rate of renal enhancement CT, YiDiXie ™-HS was applied to renal enhanced CT-negative patients.

As shown in Table 5, YiDiXie ™-HS had a sensitivity of 86.5% (95% CI: 72.0% - 94.1%) and its specificity was 84.3% (95% CI: 72.0% - 91.8%) in patients with negative renal enhanced CT. This means that the application of YiDiXie™-HS reduces the false negative rate of enhanced CT by 86.5% (95% CI: 72.0% - 94.1%).

### Diagnostic Performance of YiDiXie™-D in renal enhanced CT-positive patients

The false-positive consequences are significantly worse than the false-negative consequences in certain patients with positive enhanced CT, so YiDiXie ™-D is applied to these patients to reduce their false-positive rate.

As shown in Table 5, YiDiXie ™-D had a sensitivity of 73.9% (95% CI: 67.5% - 79.4%) and its specificity was 92.9% (95% CI: 68.5% - 99.6%) in patients with positive renal enhanced CT. This means that YiDiXie ™-D reduces the false positive rate of enhanced CT by 92.9% (95% CI: 68.5% - 99.6%).

### Diagnostic Performance of YiDiXie™-D in renal enhanced CT-negative patients

Certain enhanced CT-negative patients had significantly more severe false-positive than false-negative consequences, so the more specific YiDiXie™-D was applied to such patients.

As shown in Table 5, YiDiXie ™-D had a sensitivity of 64.9% (95% CI: 48.8% - 78.2%) and its specificity was 92.2% (95% CI: 81.5% - 96.9%) in patients with negative enhanced CT. This means that YiDiXie™-D reduces the false-negative rate of enhanced CT by 64.9% (95% CI: 48.8% - 78.2%) while maintaining high specificity.

## DISCUSSION

### Clinical significance of YiDiXie™-SS in renal enhanced CT-positive patient

The YiDiXie™test consists of three tests with very different characteristics: YiDiXie™-HS, YiDiXie™-SS and YiDiXie ™-D^13^. YiDiXie ™-HS balances sensitivity and specificity with high sensitivity and specificity, while YiDiXie ™-SS has very high sensitivity for all types of malignant tumors, but has a slightly lower level of specificity^13^. The YiDiXie™-D has very high specificity for all malignant tumor types, but low sensitivity^13^.

In patients with positive renal ultrasound, the sensitivity and specificity of further diagnostic methods are important. The trade-off between sensitivity and specificity is essentially a trade-off between the “danger of missing malignant tumors” and the “danger of misdiagnosing benign tumors”. Usually, if a benign kidney tumor is misdiagnosed as a malignant tumor, it will usually likely lead to unnecessary surgery, but it will not affect the patient’s prognosis, and its treatment cost is much lower than that of advanced cancers.Moreover, there is a higher positive predictive value in the renal enhanced CT-positive patient. It can be more harmful when the false negative rate is equal to the false positive rate. Therefore, YiDiXie ™-SS, which has a very high sensitivity but a slightly lower specificity, was chosen to reduce the false-positive rate of renal enhanced CT.

As shown in Table 5, YiDiXie ™-SS had a sensitivity of 98.6% (95% CI: 95.8% - 99.6%) in renal enhanced CT-positive patients and its specificity was 71.4% (95% CI: 45.4% - 88.3%). The above results suggest that YiDiXie ™-SS reduces the false-positive rate of renal-enhanced CT by 71.4% (95% CI: 45.4% - 88.3%) while maintaining a sensitivity close to 100%.

The above results imply that YiDiXie ™-SS substantially reduces the probability of erroneous surgery for benign renal tumors without essentially increasing the number of malignant tumors that are underdiagnosed. In other words, YiDiXie ™-SS substantially reduces the mental suffering, expensive examination and surgical costs, radiological injuries, surgical injuries, and other undesirable consequences for patients with false-positive enhanced CT of renal tumors in the case of basically no increase in delayed treatment of malignant tumors. Therefore, YiDiXie ™-SS satisfies the clinical needs well and has important clinical significance and wide application prospects.

### Clinical significance of YiDiXie™-HS in renal enhanced CT-negative patients

For patients with negative enhanced CT, both the sensitivity and specificity of further diagnostic methods are important. To balance both the sensitivity and specificity is essentially to balance the “danger of underdiagnosis of malignant tumors” and the “danger of misdiagnosis of benign tumors”. Higher false-negative rates mean that more malignant tumors are underdiagnosed, leading to delayed treatment and progression of malignant tumors, which may even develop into advanced stages. As a result, patients will have to bear the adverse consequences of poor prognosis, short survival, poor quality of life, and high cost of treatment.

In general, when benign renal tumors are misdiagnosed as malignant tumors, they usually undergo tumor resection or radical resection, which does not affect the patient’s prognosis, and their treatment costs are much lower than those of advanced cancers. Therefore, in patients with negative enhanced CT, the “risk of malignant tumor misdiagnosis” is higher than the “risk of benign tumor misdiagnosis”. Therefore, YiDiXie™-HS, with its high sensitivity and specificity, was chosen to reduce the false-negative rate of enhanced CT for renal tumors.

As shown in Table 5, the sensitivity of YiDiXie™-HS in renal enhanced CT-negative patient was 86.5% (95% CI: 72.0% - 94.1%) and its specificity was 84.3% (95% CI: 72.0% - 91.8%). These results suggest that the application of YiDiXie ™-HS reduced the false-negative rate of enhanced CT by 86.5% (95% CI: 72.0% - 94.1%).

The above results imply that YiDiXie ™-HS substantially reduces the probability of missed malignant tumors with negative enhancement CT. In other words, YiDiXie™-HS substantially reduces the poor prognosis, high treatment cost, poor quality of life, and short survival of patients with false-negative enhanced CT. Therefore, YiDiXie ™-HS fulfills the clinical needs well and has important clinical significance and wide application prospects.

### Clinical significance of YiDiXie™-D

Renal tumors considered malignant usually receive surgical treatment. However, there are some conditions that require extra caution in choosing whether to operate or not, hence further diagnosis, e.g., smaller tumors, tumors with difficulty in preserving the kidney, tumors requiring radical surgery, tumors in isolated kidneys, contralateral renal insufficiency, total renal insufficiency, and patients in poor general condition.

In patients with renal tumors, the sensitivity and specificity of further diagnostic methods are both critical. Trading off both the sensitivity and specificity is essentially a trade-off between the “danger of malignant tumors being missed” and the “danger of benign tumors being misdiagnosed”. Since smaller tumors have a lower risk of tumor progression and distant metastasis, the “risk of malignant tumor underdiagnosis” is much lower than the “risk of benign tumor misdiagnosis”. As for tumors with difficulty in preserving kidney and tumors requiring radical surgery, because the affected side of the kidney needs to be removed, the “risk of misdiagnosis of benign tumors” is much higher than the “risk of malignant tumor misdiagnosis”. In patients with isolated kidney, contralateral renal insufficiency or total renal insufficiency, the risk of renal insufficiency or even the need for dialysis after surgery is higher, so the “harm of misdiagnosis of benign tumors” is much higher than the “harm of malignant tumor underdiagnosis”. As for patients with poor general conditions, the risk of misdiagnosis of benign tumors is much higher than that of malignant tumors because the perioperative risk is much higher than that of general conditions. Therefore, for these patients, YiDiXie ™-D, which has high specificity but slightly lower sensitivity, was chosen to reduce the false-positive rate of renal-enhanced CT or to significantly reduce its false-negative rate while maintaining high specificity.

As shown in Table 5, the sensitivity of YiDiXie™-D in renal enhanced CT-positive patients was 73.9% (95% CI: 67.5% - 79.4%), and its specificity was 92.9% (95% CI: 68.5% - 99.6%); and that of YiDiXie™-D in enhanced CT-negative patients was 64.9% (95% CI: 48.8% - 78.2%). 78.2%), and its specificity was 92.2% (95% CI: 81.5% - 96.9%). These results suggest that YiDiXie™-D reduces the false-positive rate of enhanced CT by 92.9% (95% CI: 68.5% - 99.6%) or reduces the false-negative rate of enhanced CT by 64.9% (95% CI: 48.8% - 78.2%) while maintaining a high specificity.

The above results imply that YiDiXie ™-D substantially reduces the probability of wrong surgery for these patients that require additional caution in surgery. In other words, YiDiXie ™-D substantially reduces the risk of adverse outcomes such as surgical trauma, organ removal, renal insufficiency, renal dialysis, and even death and other serious perioperative complications in these patients. Therefore, YiDiXie™-D satisfies the clinical needs well and has important clinical significance and wide application prospects.

### YiDiXie™test has the potential to solve two challenges of renal tumor

First of all, the 3 tests of the YiDiXie™test are of clinical importance in renal tumors. As mentioned earlier, YiDiXie™-SS, YiDiXie™-HS and YiDiXie ™-D have significant diagnostic value in patients with positive or negative enhanced CT, respectively.

Second, the 3 tests of YiDiXie ™test can significantly relieve clinicians’ work pressure and facilitate on-time diagnosis and timely treatment of malignant tumor cases that would otherwise be delayed. On the one hand, YiDiXie™-SS can relieve surgeons of non-essential work significantly. Patients with renal tumors that are positive on enhanced CT usually undergo surgery. The timely completion of these surgeries is directly dependent on the number of surgeons. In many parts of the world, appointments are booked for months or even more than a year. This inevitably delays the treatment of malignant cases among them, and thus it is not uncommon for patients with renal tumors awaiting surgery to develop malignant progression or even distant metastases. As shown in Table 5, YiDiXie™-SS reduced the false-positive rate of renal enhanced CT by 71.4% (95% CI: 45.4% - 88.3%) with essentially no increase in renal cancer missed diagnosis. As a result, YiDiXie ™-SS can greatly relieve the stress of non-essential work for surgeons, facilitating timely diagnosis and treatment of renal tumors or other diseases that would otherwise be delayed.

On the other hand, YiDiXie™-HS and YiDiXie™-D can greatly relieve clinicians’ work pressure. In cases of difficult diagnosis by enhanced CT, this patient usually requires an enhanced MRI or puncture biopsy. The timely completion of these enhanced MRIs or renal puncture biopsies is directly dependent on the number of clinicians available. Appointments are available for months or even more than a year in many areas of the world. It is not uncommon for patients with renal tumors waiting for an enhanced MRI or puncture biopsy to experience malignant progression or even distant metastasis. YiDiXie ™-HS and YiDiXie ™-D can be used as alternatives to these enhanced MRIs or puncture biopsies, greatly relieving clinicians of their workloads and facilitating timely diagnosis and treatment of other tumors that would otherwise be delayed.

Final, the YiDiXie™test enables “just-in-time” diagnosis of renal tumors. On the one hand, the YiDiXie ™test requires only microscopic amounts of blood, allowing patients to complete the diagnostic process non-invasively without having to leave their homes. A single YiDiXie ™test requires only 20 microliters of serum, which is equivalent to the volume of 1 drop of whole blood (1 drop of whole blood is about 50 microliters, which produces 20-25 microliters of serum)^13^. Considering the pre-test sample quality assessment experiments and 2-3 repeat experiments, 0.2 ml of whole blood is sufficient to complete the YiDiXie™test^13^. The 0.2 ml of finger blood can be collected at home using a finger blood collection needle, eliminating the need for venous blood collection by medical personnel and allowing patients to complete the diagnostic process non-invasively without having to leave their homes^13^.

On the other hand, the diagnostic capacity of the YiDiXie™test is nearly limitless. Figure 1 shows the basic flow chart of the YiDiXie ™test, which shows that the YiDiXie ™test requires neither a doctor or medical equipment, nor medical personnel to collect blood. Therefore, the YiDiXie™test is completely independent of the number of medical personnel and medical facilities, and its testing capacity is nearly unlimited. Thus, the YiDiXie ™test enables “just-in-time” diagnosis of renal tumors without the patient having to wait anxiously for an appointment.

**Figure 1.**
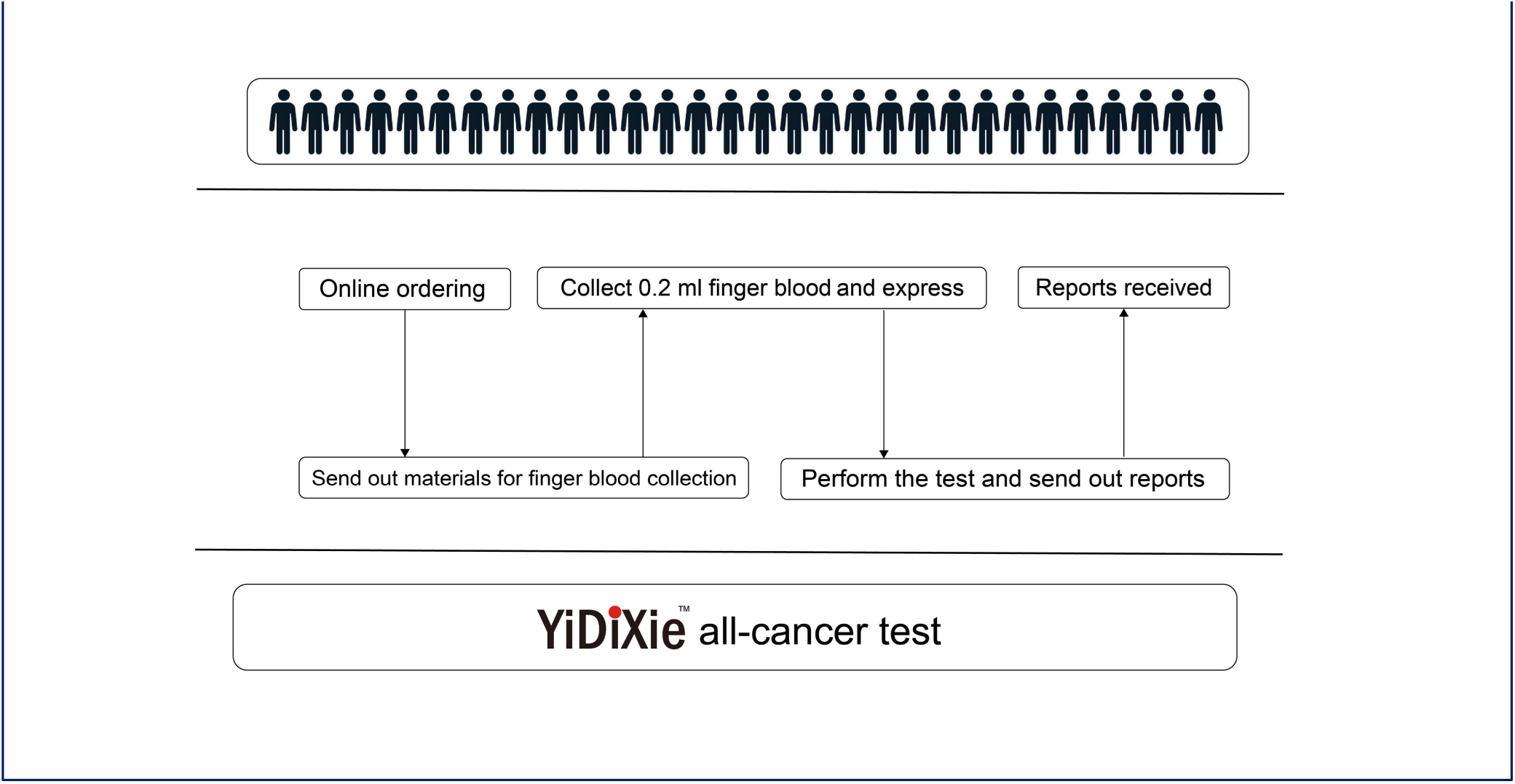
Basic flowchart of the YiDiXie™ test.

In short, YiDiXie ™test has an significant diagnostic value in renal cancer, and is expected to solve the problems of “ high false-positive rate of renal enhanced CT” and “high false-negative rate of renal enhanced CT”.

### Limitations of the study

First, the number of cases in this study was relatively small, and larger sample size clinical studies are needed for further evaluation in the future.

Second, this study was a malignant tumor case-benign tumor control study in hospitalized patients, and a cohort study in the natural population of renal tumors is needed for further evaluation in the future.

Last, this study was a single-center study, which may have led to some degree of bias in the results of this study. A multi-center study is needed in the future to further assess the results.

## CONCLUSION

YiDiXie™-SS has very high sensitivity and high specificity in renal tumors. YiDiXie ™-HS has high sensitivity and high specificity in renal tumors. YiDiXie ™-D has high sensitivity and very high specificity in renal tumors. YiDiXie™-SS significantly reduces renal enhancement CT false-positive rates with essentially no increase in delayed treatment of malignant tumors. YiDiXie ™ -HS significantly reduces the false-negative rate of renal enhancement CT. YiDiXie ™-D can significantly reduce the false-positive rate of renal enhancement CT or significantly reduce the false-negative rate while maintaining high specificity. the YiDiXie ™test has important diagnostic value in renal cancer and is expected to solve the two problems of “too high false-positive rate” and “too high false-negative rate” of renal enhanced CT.

## Data Availability

All data produced in the present study are contained in the manuscript.

## REFERENCES

1. Motzer R J, Jonasch E, Agarwal N, et al. Kidney Cancer, Version 3.2022, NCCN Clinical Practice Guidelines in Oncology [J]. Journal of the National Comprehensive Cancer Network, 2022, 20(1):71–90.

2. Bray F, Laversanne M, Sung H, et al. Global cancer statistics 2022: GLOBOCAN estimates of incidence and mortality worldwide for 36 cancers in 185 countries [J]. CA Cancer J Clin, 2024.

3. Sung H, Ferlay J, Siegel R L, et al. Global Cancer Statistics 2020: GLOBOCAN Estimates of Incidence and Mortality Worldwide for 36 Cancers in 185 Countries [J]. CA: A Cancer Journal for Clinicians, 2021, 71(3):209–49.

4. Ljungberg B, Albiges L, Abu-Ghanem Y, et al. European Association of Urology Guidelines on Renal Cell Carcinoma: The 2022 Update [J]. European Urology, 2022, 82(4):399–410.

5. Bukowski R M. Natural history and therapy of metastatic renal cell carcinoma [J]. Cancer, 1997, 80(7):1198–220.

6. Network NCC. NCCN Clinical Practice Guidelines in Oncology Kidney Cancer Version 4.2024 [J]. 2024.

7. Capitanio U, Bensalah K, Bex A, et al. Epidemiology of Renal Cell Carcinoma [J]. European Urology, 2019, 75(1):74–84.

8. Stephenson A J, Kuritzky L, Campbell S C. Screening for urologic malignancies in primary care: Pros, cons, and recommendations [J]. CLEVELAND CLINIC JOURNAL OF MEDICINE, 2007, 74.

9. Corcoran A T, Russo P, Lowrance W T, et al. A Review of Contemporary Data on Surgically Resected Renal Masses—Benign or Malignant? [J]. Urology, 2013, 81(4):707–13.

10. Borghesi M, Brunocilla E, Volpe A, et al. Active surveillance for clinically localized renal tumors: An updated review of current indications and clinical outcomes [J]. International Journal of Urology, 2015, 22(5):432–8.

11. Kim J H, Sun H Y, Hwang J, et al. Diagnostic accuracy of contrast-enhanced computed tomography and contrast-enhanced magnetic resonance imaging of small renal masses in real practice: sensitivity and specificity according to subjective radiologic interpretation [J]. World Journal of Surgical Oncology, 2016, 14(1).

12. Wildberger J E, Adam G, Boeckmann W, et al. Computed Tomography Characterization of Renal Cell Tumors in Correlation with Histopathology [J]. Investigative Radiology, 1997, 32(10):596–601.

13. Chen Sun, Chong Lu, Yongjian Zhang, et al. Evaluation of the Multi-Cancer Early Detection (MCED) value of YiDiXie™-HS and YiDiXie™-SS [J]. medRxiv, 2024:doi: 10.1101/2024.03.11.24303683.

14. Edge SB and Compton CC: The American Joint Committee on Cancer: the 7th Edition of the AJCC Cancer Staging Manual and the Future of TNM. Annals of Surgical Oncology. 17: 1471–1474, 2010.

15. Amin MB, Greene FL, Edge SB, Compton CC, Gershenwald JE, Brookland RK, Meyer L, Gress DM, Byrd DR and Winchester DP: The Eighth Edition AJCC Cancer Staging Manual: Continuing to build a bridge from a population-based to a more “personalized” approach to cancer staging. CA: A Cancer Journal for Clinicians. 67: 93–99, 2017.

